# Changes of sociobehavioural characteristics and HIV in 29 Sub-Saharan African countries, 2000-2018

**DOI:** 10.1101/2023.02.16.23286024

**Authors:** Jeffrey Post, Wingston Ng’ambi, Olivia Keiser, Aziza Merzouki

## Abstract

**Introduction:** HIV epidemics and associated sociobehavioural characteristics vary widely across Sub-Saharan African (SSA) countries. In this study, we investigated how national changes on sociobehavioural characteristics may be associated with trajectories in HIV epidemics.

**Methods:** We analyzed aggregated data from 80 DHS surveys of 29 SSA countries between 2000 and 2018. We described each national survey by 46 pre-selected demographic, socio- economic, behavioural and HIV-related variables. Using Principal Component Analysis, we reduced the dimensionality of surveys and visualized them on a 2-dimensional space. We used consensus clustering to identify groups of countries with similar sociobehavioural characteristics over time, and identified the main drivers of change. We used k-means clustering with dynamic time warping to group countries based on the evolution of their national HIV effective contact rates since 1990 (computed based on UNAIDS estimates of yearly HIV incidence and prevalence).

**Results:** We identified three groups of countries with similar sociobehavioural and HIV-related characteristics that persisted from 2000 to 2018. While countries within groups remained similar since 2000, differences between the groups increased over time. The most important characteristics that explained the change in the sociobehavioural space since 2000 were: ART coverage, HIV testing, increase in accepting attitudes towards People Living with HIV/AIDS and increasing knowledge about AIDS. We also identified two groups of countries with characteristic evolutions of HIV effective contact rates since 1990. The first group of countries was characterized by a rapid decrease of effective contact rate since the early 1990. The second group experienced a slow decrease of the effective contact rate in the early 1990, followed by a faster decrease after 1995. While the three clusters based on sociobehavioural characteristics were associated with different peak levels of HIV incidence, the two clusters based on the progression of the effective contact rate were associated with the timing when peak HIV incidence levels were reached.

**Conclusions:** Our findings suggest that the initial conditions of nascent epidemics, likely determined by sociobehavioural characteristics, are crucial in determining the long-term progression of HIV epidemics. Our methods can help to predict the effects of behavioural change on the HIV epidemic and design targeted interventions to reach epidemic control.

## Introduction

Sub-Saharan Africa (SSA) accounts for 60% of the 4000 daily new infections that occur globally, and accounted for 67% of the 680’000 AIDS related deaths in 2020(1). Levels of HIV epidemics vary widely between and within countries, and this heterogeneity may be largely explained by diverse cultural and sociobehavioural characteristics. Thirty-five years since the World Health Organization (WHO) has launched the Special Program on AIDS, renewed and sustained action is needed to get a better understanding of the sociobehavioural drivers of the epidemic and to get closer to “ending the AIDS epidemic as a public health threat”(2).

Previous studies that analyzed socio-behavioral risk factors of HIV were generally cross-sectional and with narrow scopes, limited to single countries(3), single regions (4), or specific key populations (5,6). Other studies had a wider geographic scope, but they analyzed only few risk factors (7). More recent studies analyzed changes of some HIV risk factors and behaviors over time(8–10). These studies often used descriptive statistics and regression analyses. In a previous study (11), Merzouki et al. assessed sociobehavioural heterogeneity across 29 SSA countries based on 48 demographic, socio-economic, behavioural and HIV-related attributes and their relation to HIV incidence using clustering techniques, but the study was cross-sectional only.

In the pursuit of HIV epidemic control, numerous programs and interventions have been implemented. It has become increasingly complex to gauge the impact of interventions, all while becoming ever more important to prioritize efforts (12). Most previous studies used HIV incidence as a standard metric to gauge the various risk factors and outcomes, but this may fall short in a context of changing HIV/AIDS epidemiological transition from a pandemic to low level endemic disease (13).

In this study we analyzed sociobehavioural data aggregated at the national-level for 29 SSA countries. We used unsupervised machine learning techniques to identify major sociobehavioural trends and assess heterogeneity across SSA, both spatially and across time. In parallel, we assessed the progression of the national HIV epidemics using the effective contact rate.

## Methods

### Data

We analyzed data from the Demographic Health Surveys (DHS). The DHS has been gathering nationally representative data at regular intervals since its inception in 1984 (14). The data span social, behavioral, geographic, economic, and health related issues, including HIV-related data and sexual behavior, especially since the emergence of HIV/AIDS as a public health crisis (15).

We intended to include all indicators possibly influencing the risk of acquiring HIV for individuals aged 15-49 years old, or possibly affecting the course of HIV epidemics on local, regional, and national levels. We used the DHS program, STATcompiler (16), to collect the aggregated data from surveys of 29 SSA countries between 2000 and 2019, that were available up to September 2020. While we aimed to go as far back in time as possible to include the emergence of HIV, earlier surveys lacked specific indicators of interest, resulting in a total of 80 surveys collected, with each of the 29 countries having between 1 and 6 surveys over that time period; see Table 1.

**Table 1.**
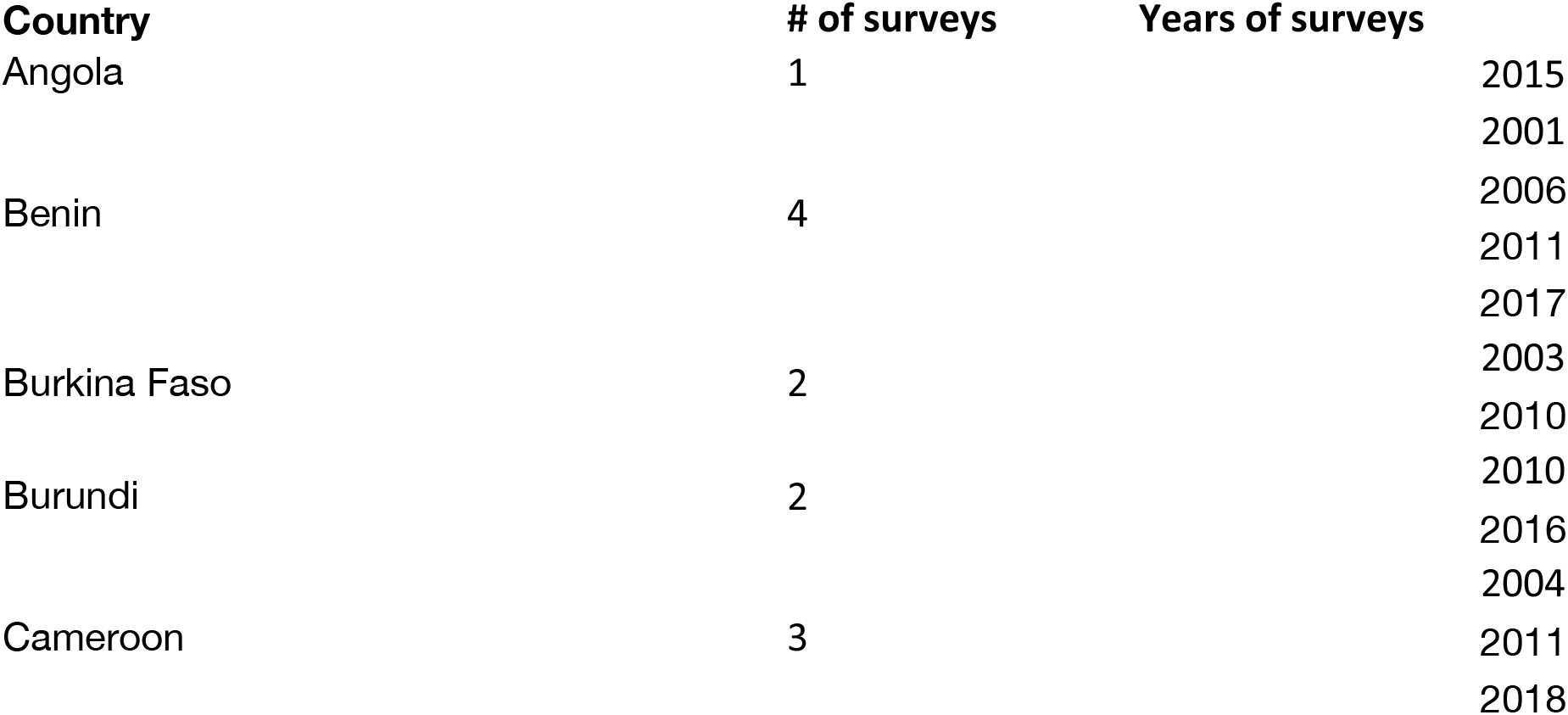
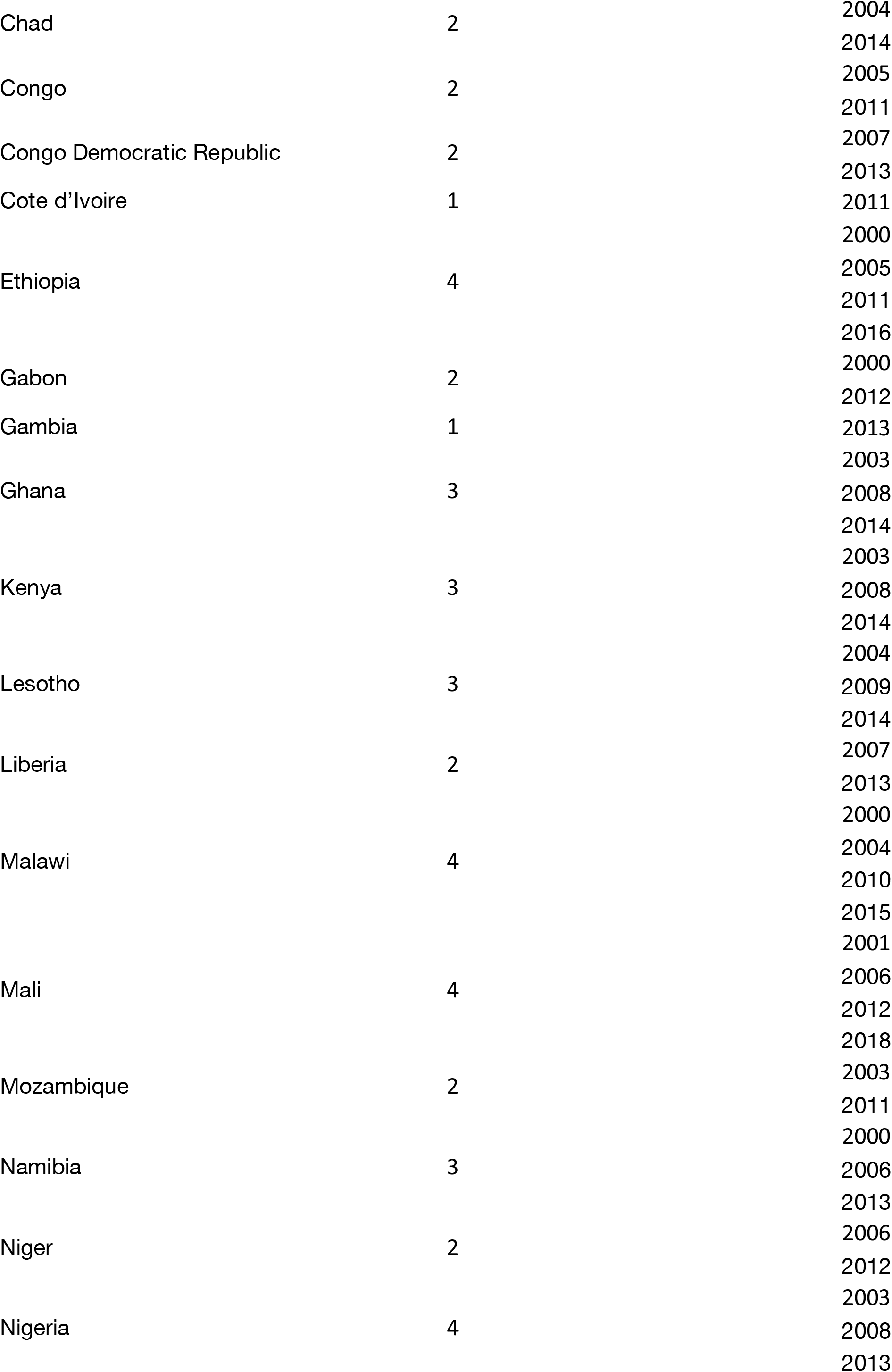
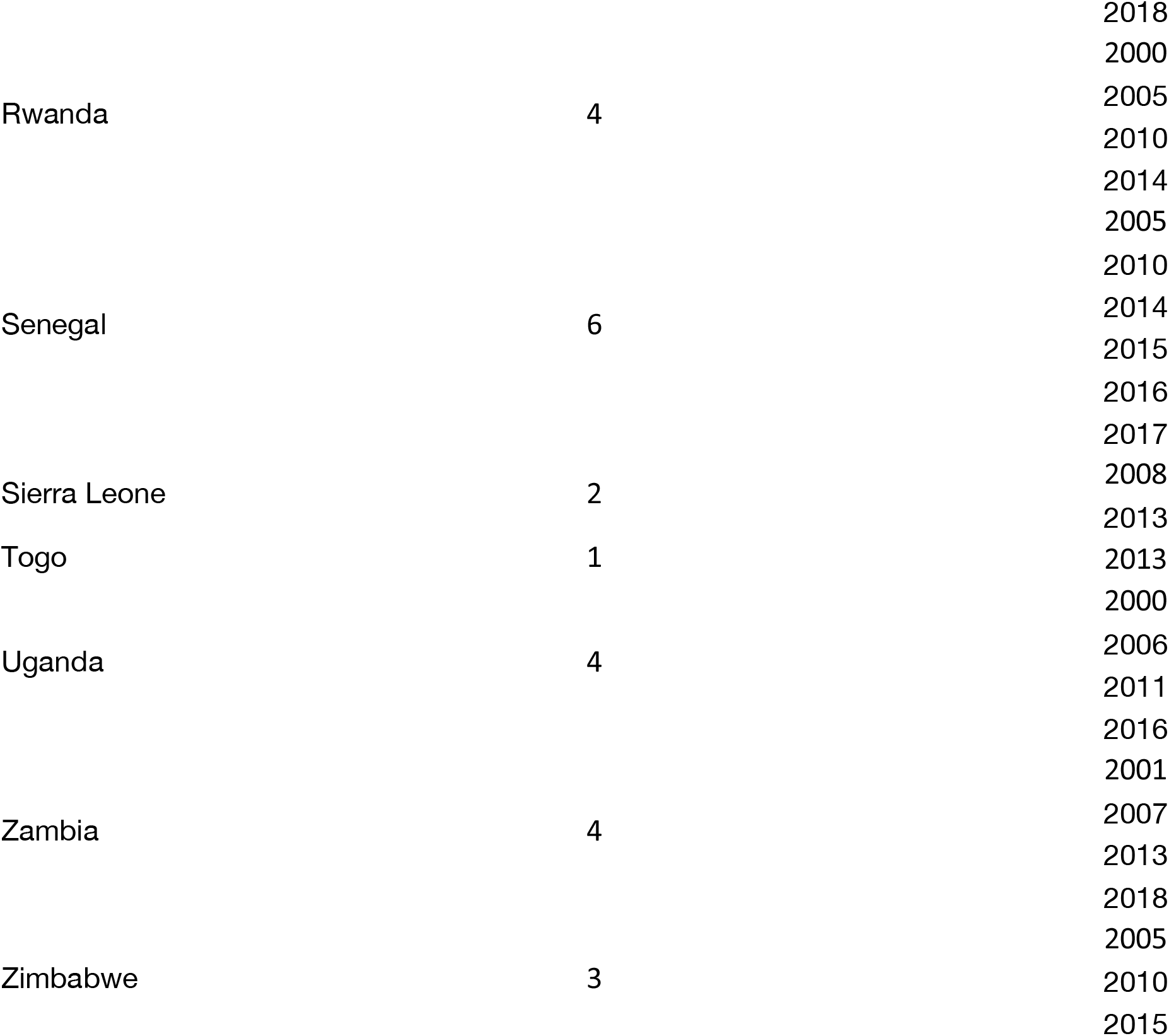
List of countries with number and years of surveys

As in a previous publication (11), we retained only variables that varied significantly across regions and did not strongly correlate with other variables (Table S1 in Supplementary Material). The data were represented as percentages, with mean number of sexual partners in lifetime scaled using min-max normalization. The resulting 46 attributes create a 46- dimensional image of each survey.

While most of these attributes were collected using the DHS program, STATCompiler (16), we also used additional data from the Institute of Health Metrics and Evaluation for male circumcision data(17), the World Bank for urban/rural divide data(18), Gini wealth coefficient(19) and ART coverage (20), and The Correlates of War Project for religious data (21). Attributes with missing variables for which alternative sources were unavailable were imputed using additional topic related variables available from the DHS using multiple iterative chained equations (see Supplementary Material). Finally, estimates for national HIV incidence and prevalence between the years 1990 and 2019 were obtained from the latest (2020) UNAIDS’ AIDSinfo (22).

### Analysis

#### Trends of sociobehavioural characteristics

We reduced the dimensionality of each survey using Principal Component Analysis (PCA) (23) (See details in Supplementary Material). To capture the trends in our dataset, and to facilitate comparison with previous results(11), we computed the rotation matrix using the most recent surveys collected since 2015. This rotation matrix was then used to project all other surveys onto the dimensionally reduced PCA space. We identified the trends of all national surveys by both visualizing and quantifying their trajectories on the 2D PCA space and pointed to the key sociobehavioural characteristics driving these changes.

To capture sociobehavioural heterogeneity across SSA, we used a consensus clustering method to identify groups of similar surveys in terms of sociobehavioural characteristics. We identified an optimum number of three clusters by maximizing the Silhouette score(11). The consensus clustering method aggregated results from three hierarchical clusterings and three k-means clusterings (24), each run on a different number of PCs. The first hierarchical and k-means clusterings run with two main PCs only; the second clusterings run with the *n* PCs that accounted for 95% of the overall variance; and the third run with the entire 46-dimensional data set. Thus, we were able to construe sociobehavioural heterogeneity and homogeneity across SSA, both spatially and over time.

### Progression of HIV effective contact rate over time

We calculated for each country a yearly effective contact rate β from 1990 to 2019 based on estimated HIV incidence and prevalence (see Supplementary Material). The effective contact rate β is a measure of incidence given the prevalence (i.e. the number of new infections given the local context of the HIV epidemic) and is effectively both a proxy for the sociobehavioural characteristics of the population while also directly gauging the transmission dynamics of HIV within that population, doing so independently of the local context of the HIV epidemic. We compared the progression of the effective contact rates (ECR) since 1990 and since 2000 (as the DHS data only goes back that far) across SSA to that of HIV incidence and compared them across the clusters obtained previously and visualized them with box plots.

We identified clusters of countries based on the evolution of their ECR by first scaling the time-series such that they had zero mean and unit variance (following the assumption that their amplitudes are not as informative as their relative shapes) and used k-means clustering (24) with a dynamic time warping barycenter averaging algorithm (25). We measured the quality of the clustering results using a silhouette score (26) and determined the optimal number of clusters, which maximized that score. We compared this new set of clusters with the ones previously obtained using sociobehavioural characteristics and compared their respective HIV incidence and ECR.

### Tools and software

We used the open source Python language (27), Pandas (28), NumPy(29), SciPy (30), TSLearn (31), Scikit-learn (32) for data aggregation, wrangling, and analysis, and used Plotly (33) for data visualizations.

### Ethics approval

This study used only publicly accessible aggregate-level data. No ethics approval was needed.

## Results

We analyzed 80 surveys from 29 SSA countries between the years 2000 and 2018, see Table 1. HIV incidence ranged from 0.09 new cases per 1000 population in Niger in 2018 to 50.04 new cases per 1000 population in Zimbabwe in 1991. HIV prevalence ranged from less than 0.1% in Gambia and Senegal in 1990, to 25.4% in Zimbabwe in 1996. HIV mortality ranged from 0.01 per 1000 population in Benin in 1990 to 12.83 per 1000 population in Zimbabwe in 2003. Likewise, sociobehavioural characteristics varied substantially across SSA countries and surveys (see Table S2 in Supplementary Material).

### Trends of sociobehavioural characteristics

Using PCA we found that 95% of the variance in the original 46-dimensional data set can be explained by the first 8 PCs, with the first (PC-1) and the second (PC-2) explaining 60.7% and 12.5% respectively of the total variance across those surveys done in SSA after 2015 (Figure S1 in Supplementary Materials).

Projecting the SSA surveys since 2015 onto the 2D PCA space allows us to identify three distinct clusters of countries that form an upside-down V-shape (Figure 1 - top). The variable correlation plot (Figure 1 - bottom) highlights that countries of the first (red) cluster on the lower left-hand side are characterized by more accepting attitudes towards PLWHA, better knowledge of HIV, higher literacy rates, higher ART coverage and HIV testing, but notably lower rates of male circumcision. Similarly, countries of the second (yellow) cluster on the lower right-hand side, are characterized by a high percentage of Muslim populations, lower mean number of sexual partners, and fewer women participating in decisions. Countries of the third (orange) cluster, higher on the PC-2 axis are characterized by a less rural population, stronger women empowerment (women who work and disagree with domestic violence), and high rates of male circumcision. There is a geographical component to the clustering as shown in Figure S2 (Supplementary Material) with the first cluster being composed mostly of countries of Southern and Eastern Africa (Burundi, Kenya, Lesotho, Malawi, Rwanda, Uganda, Zambia, and Zimbabwe), the second cluster of countries of the Sahel region (Chad, Mali, and Senegal), and the third of countries of mostly Western and Central Africa (Angola, Benin, Cameroon, Ethiopia, Ghana, and Nigeria).

**Figure 1.**
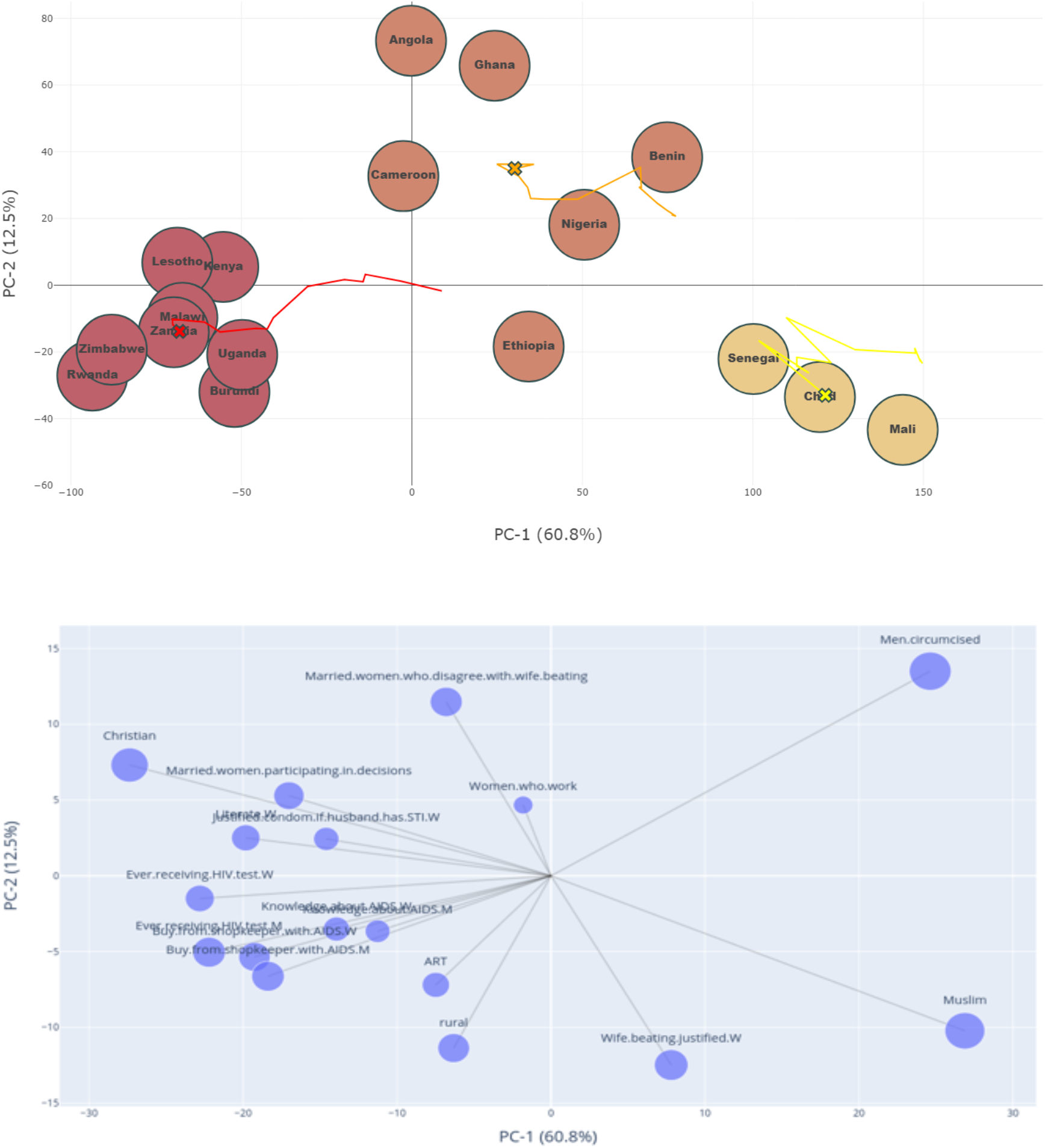
Projection of countries (top), and original sociobehavioural indicators (bottom) onto the reduced 2-dimensional space given by PC-1 and PC-2. The size of the dots on the bottom panel represent their contribution to PC1 and PC2 (in %).

Projecting earlier surveys onto the same 2D-space gives an insight into the evolution of sociobehavioural characteristics in SSA from 2000 to 2018 (Figure 2). The reversed V-shape was mostly preserved through time, and although there are some edge cases like Ethiopia (between Sahel and western/central countries) and Gabon (between western/central and eastern/southern countries), the geographical and sociobehavioural similarities remain strong and the clusters easily distinguishable. From 2000 to 2018 we found the first cluster to move the most (∼85 units in the negative PC-1 direction; 20 units in the negative PC-2 direction), and the second cluster drifting the least (only 30 units in the negative PC-1 direction; 10 units in the negative PC-2 direction). The sociobehavioural heterogeneity across clusters tended to increase over time as measured by the 2D Cartesian distance between them; from 2000 to 2018, the first and second clusters moved from 142 to 190 units away, the first and third clusters moved from 70 to 110 units, and the second and third clusters moved from 90 to 110 units.

**Figure 2.**
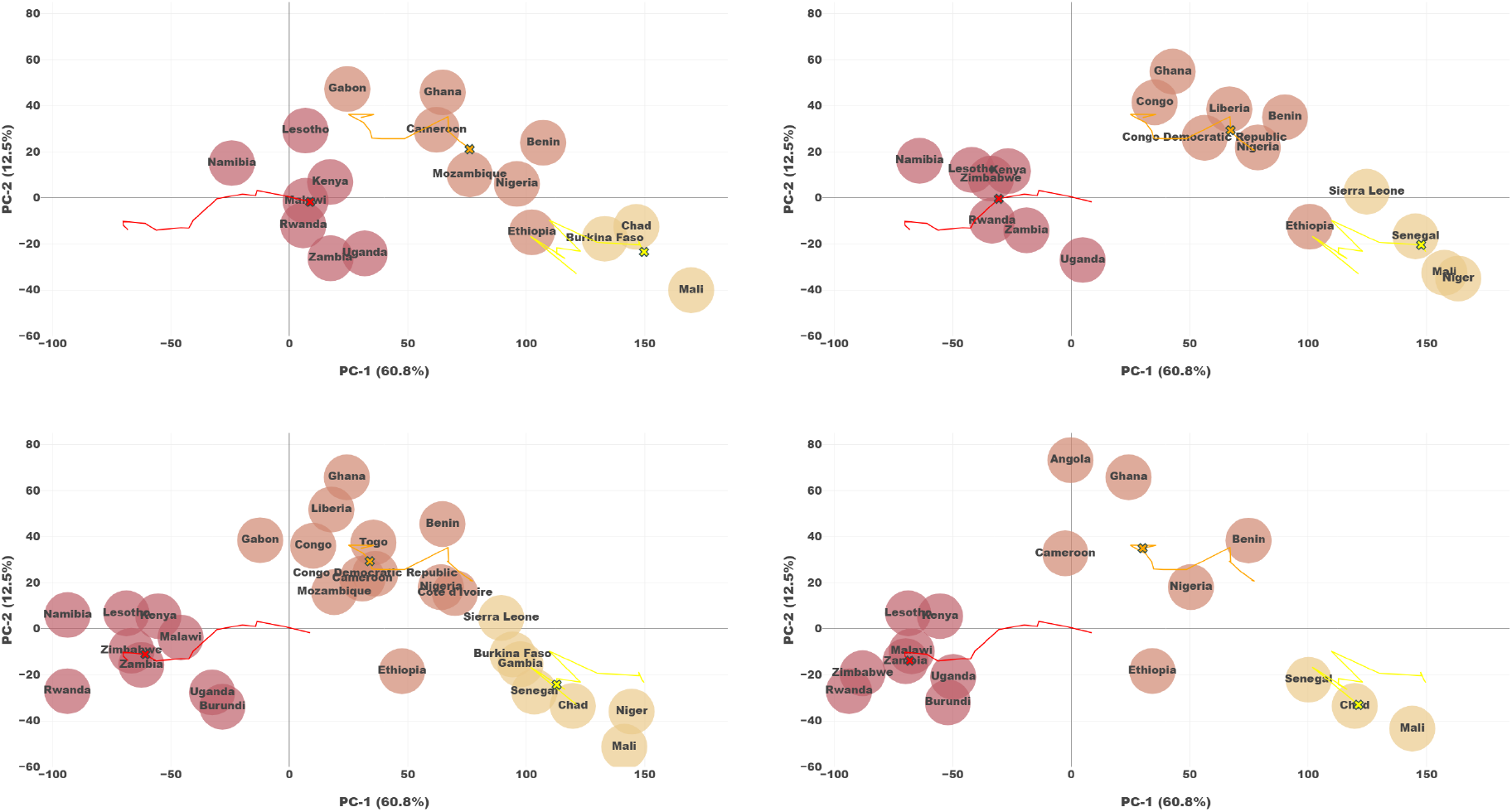
Visualization of progression of countries on the 2-dimensional space given by PC1- and PC-2. Color represents sociobehavioural (SB) cluster - Lines represent trajectory of cluster centroids over 2000-2018 with current position marked with a circle. (a) Surveys gathered between the years 2000 and 2004. (b) Surveys gathered between the years 2005 and 2009. (c) Surveys gathered between the years 2010 and 2014. (d) Surveys gathered since 2015.

Since 2000 there has been a clear, though varying, shift of the three clusters towards the negative PC-1 direction. This shift corresponds in all three clusters to an increase in HIV testing, particularly for women (increase from 15% to 85% for the first cluster, 5% to 42% for the second cluster, and 7% to 46% for the third cluster), an increase in ART coverage (increasing to 60%, 42%, and 54% coverage in the first, second, and third clusters respectively since its introduction in the early 2000s), and an increase in acceptance of PLWHA (51% to 80% for the first cluster, 26% to 45% for the second cluster, and 27% to 50% for the third cluster) (Figure 3). While this leftwards shift is slowed in the second cluster (countries of Sahel region) by a decrease in working women (70% to 50%), the trend is accentuated in the other two clusters by increases in knowledge about HIV (28% to 51% for the first cluster and 18% to 30% for the third cluster), increases in women empowerment (women participating in decision making and disagreeing with domestic violence), and an increase in working men for the first cluster in red. Movement on the PC-2 axis is smaller and it is difficult to discern an overall trend. While the strong increase in ART coverage across the three clusters tends to push them in the negative PC-2 direction, the third cluster experiences higher increases in women empowerment (married women who disagree with wife beating and rates of working women) which can explain the positive movement on the PC-2 axis despite the gains made in ART coverage. While significant changes were observed during the last two decades in uptake of HIV testing, acceptance of PLWHA, knowledge about AIDS, ART coverage, other HIV risk factors such as the rate of male circumcision remained relatively constant across the three SB clusters (Figure S3 in Supplementary Material).

**Figure 3.**
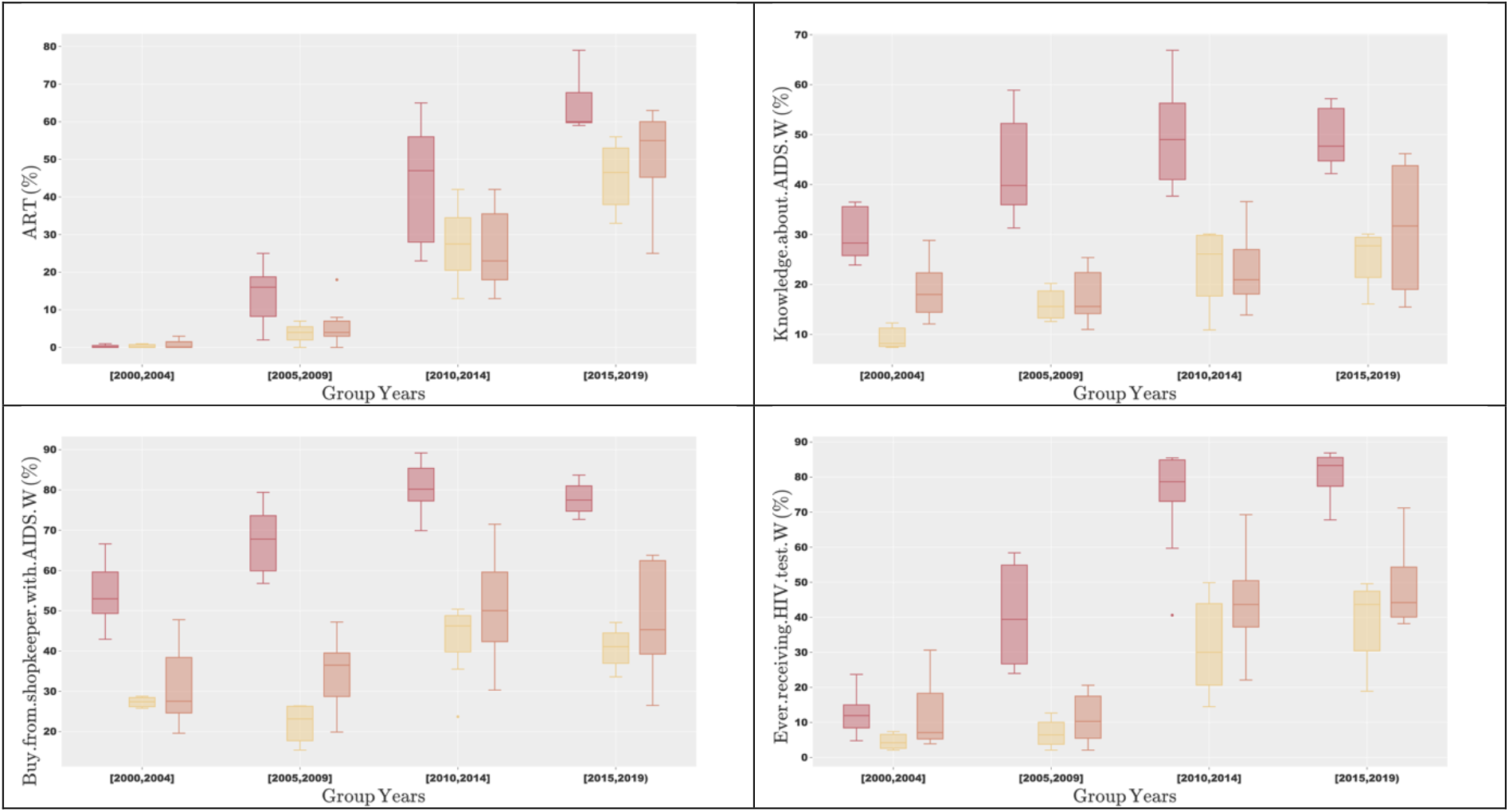
Progression of HIV-related attributes between 2000 and 2018. Colors represent SB clusters. (a) Progression of ART coverage. (b) Progression of knowledge about AIDS (women). (c) Progression of attitudes towards PLWHA (women). (d) Progression of women who ever received an HIV test.

### Progression of HIV effective contact rate β over time

Figure S4 (in Supplementary Material) shows the progression of the ECR across SSA since 1990. While there was a sharp decrease from 1990 to 2000, with substantial variation across SSA in 1990 to a point where low values are standard across SSA in 2019, we found that the three clusters had a remarkably similar progression of ECR. We did notice a slightly higher average initial value of ECR for countries of the first cluster compared to the other two clusters (0.30 compared to 0.27 and 0.26 for countries of Sahel region and countries of central/western SSA respectively). In contrast, in 2019, countries of the first cluster tend to have a slightly lower average value of β when compared to the other two clusters (0.044 on average compared to 0.052 and 0.054 for countries of western/central SSA and countries of Sahel region respectively).

The silhouette score indicates countries of SSA formed two clusters based on their ECR evolution. The first (green) cluster includes countries whose ECR were already decreasing in 1990 and continued doing so until rapidly reaching low value steady-states around the year 2000 (from an average value of 0.266 in 1990 to 0.094 in 2000, and finally reaching 0.046 in 2019). The second group of countries (in purple) are countries whose ECR tended to decrease later around 1995, and reached low steady state values between 2005 and 2010 (an average of 0.296 in 1990, 0.153 in 2000, and finally 0.057 in 2019)(Figure 4 - a). We found that while the three original clusters based on sociobehavioural characteristics associated well with the amplitude of the HIV incidence peak (see Figure 2), the two new clusters based on effective contact rate were associated with the timing when peak HIV incidence levels were reached (Figure 4 – b; c).

**Figure 4.**
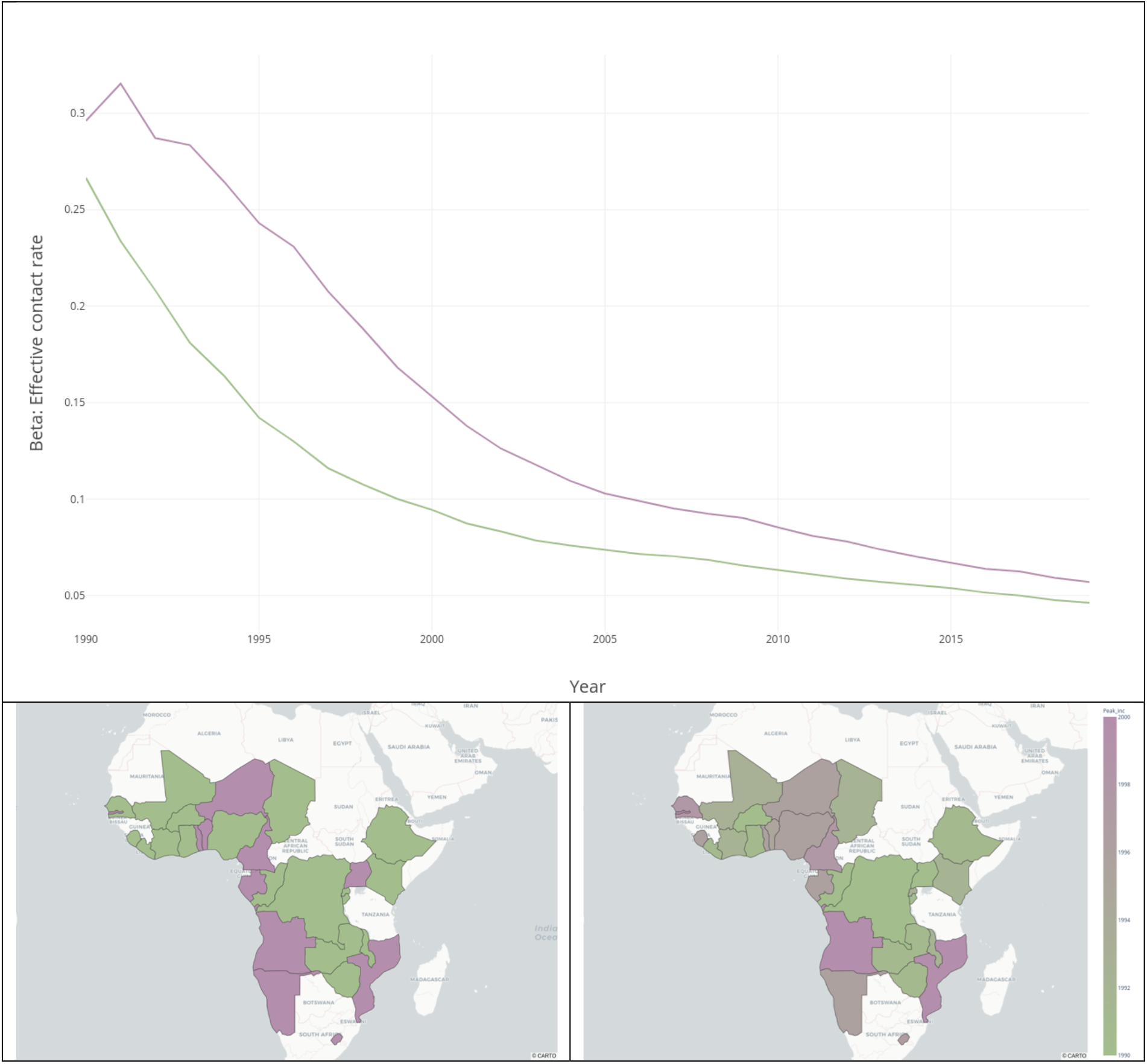
Visualization of β progression clusters across SSA. (a) Progression of effective contact rates per β progression cluster. (b) Map of SSA showing β progression clusters. (c) Map of SSA showing year of peak levels of HIV incidence.

At the country-level, we can distinguish three patterns that give insight into the relationship between evolution of the sociobehavioural characteristics and evolution of effective contact rate. Chad and Malawi (Figure S5 in Supplementary Material) exemplify the case in which two countries have different epidemiological contexts in 1990 but have very similar ECR since. In effect they are in different clusters based on sociobehavioural characteristics but in the same cluster based on effective contact rate evolution. While these two countries have markedly reduced both incidence and prevalence over this period, the relative difference that existed between the two in 1990 remains the same today. While the rate of decrease of effective contact rate somewhat slowed for both countries around the year 2000, Malawi has tended to keep decreasing at a faster rate than Chad. On the sociobehavioural space, this translates to a larger shift in the negative PC1 direction for Malawi than Chad (80 vs. 25 units), the difference stemming mostly from a much larger increase in HIV testing and increase in knowledge about HIV in Malawi than Chad. On the PC2 axis, Malawi shifts 10 units in the negative direction whereas Chad moves 20 units in the same direction - with Malawi experiencing a faster increase in women’s empowerment while Chad had a decrease in working women.

Cameroon and Ghana, on the other hand, have very similar epidemiological contexts in 1990 (incidence and prevalence of 2.2/1000 population and 0.7% for Cameroon and 2.8/1000 population and 1.3% for Ghana) but have very different evolutions of ECR from 1990 onwards (Figure S6 in Supplementary Material). This strong divergence is associated with very different evolutions of their respective HIV epidemics, with Cameroon experiencing a much higher and later peak in HIV incidence. Since around 2000 however, Cameroon has gone through a much quicker decrease of ECR when compared to Ghana, resulting in a quicker improvement of the HIV epidemic. While they remain in the same cluster based on sociobehavioural characteristics, this is seen in their different movements on the sociobehavioural space – Cameroon shifting 65 units in the negative PC1 direction while Ghana only shifted 30 units in the same negative PC1 direction. Similarly, as the previous example, the extra shift for Cameroon stems from faster increases in HIV testing and better knowledge about HIV/AIDS, coupled with a reduction in stigma associated with HIV/AIDS. On the PC2 axis, Cameroon remains at the same level over time and Ghana increases 20 units in the positive PC2 direction, likely a result of the faster uptake in ART coverage in Cameroon in recent years.

Chad and Ghana have both similar epidemiological contexts in 1990 (although they are in separate sociobehavioural clusters) and similar evolutions of ECR up to the year 2000, meaning any difference in contexts in 1990 can be seen in 2000 (Figure S7). Where they diverge however is since 2005, with Chad continuing to decrease its effective contact rate much quicker than Ghana resulting in a large difference in their current incidence and prevalence levels. We described above their movement on the sociobehavioural space, and while they move similarly on the PC1 axis, with similar increases in HIV testing and knowledge about AIDS, Chad moves in the negative direction on the PC2 axis while Ghana moves in the positive direction - this directly relates to the slower increase in coverage of ART in Ghana compared to Chad.

## Discussion

Using PCA and consensus clustering, we identified three groups of countries with similar sociobehavioural and HIV-related characteristics that persisted from 2000 to 2019. While the differences between the three groups of countries increased, countries within the same group remained similar since 2000, and the different levels of HIV incidence and prevalence present since the early 1990s across the three sociobehavioural groups have persisted over time. Using time-series K-means clustering, we also identified two distinct groups of countries with characteristic evolutions of HIV ECR between 1990 and 2019. The first group of countries was characterized by an early rapid decrease of effective contact rate, which already started in 1990, while the second group of countries experienced a slower decrease of effective contact rate in the early 1990, that accelerated after 1995. The different levels of HIV epidemics seen today in SSA countries are likely the result of the different initial sociobehavioural conditions in which the early years of the HIV epidemics took place, combined with the different evolutions of the clusters since then.

Small differences in ECR in nascent epidemics can lead to very different levels of epidemics over time (34). In the early 1990s, countries of the first cluster (eastern/southern region) had a higher effective contact rate than countries of the other two clusters (Sahel and the central/western regions). It can thus be hypothesized that the slight difference between the initial ECR of the three clusters was among the main drivers of the long-term differences over the course of the entire epidemic. Further work on reconstructing the ECR of the 1980s across SSA would allow for better insight into this hypothesis.

Male circumcision rates between the identified clusters may be one of the drivers of the early differences in ECR across clusters. Male circumcision has been shown to have a protective effect for HIV acquisition for seronegative men (35–37). This could in turn reduce the ECR, all else being equal, of countries with high rates of male circumcision versus countries with low rates of male circumcision. Countries from the second and third SB clusters with high rates of male circumcision (Median (IQR) of 91.5 (95.7-93.4) and 91.5 (97.1-95.6), respectively) had on average slightly lower values of ECR when compared to countries of the third cluster with low rates of male circumcision (Median (IQR) of 15.0 (33.6-23.0)) between the years 1990 and 2000. It should be noted that although high rates of male circumcision might have had a protective effect and reduced ECR in nascent HIV epidemics, it is unclear whether voluntary medical male circumcision (VMMC) would have as much of an effect in later stages of epidemics where the ECR have already been drastically lowered (34). VMMC is a prevention priority of PEPFAR and has been an official recommendation of the WHO since 2007 (38).

While specific indicators such as circumcision rates partly explain the different effective contact rate levels across SSA in 1990, political instability in some sub-Saharan African countries coupled with an increase in rural to urban migration may have exacerbated it (39,40). On the other hand, effective political will and early recognition of HIV/AIDS as a public health threat, despite the already strong stigma associated with the disease, can explain the different timings seen for the decrease in ECR across SSA. Uganda is a prime example as it implemented the AIDS Control Program (ACP) in 1986 (41), the only such program at that time, and had the lowest effective contact rate in 1990.

Since 2005, SSA has also seen a large increase in coverage of ART with countries of the first cluster (eastern/southern SSA) reaching 81% coverage while countries of the other two clusters increased to 52% coverage on average since its introduction in the early 2000s. With high ART coverage being associated with declines in both risk of HIV acquisition and HIV mortality (42), countries of the first cluster would be expected to see a quicker decrease in ECR relative to countries of the other two clusters. Evidence of this can be seen in the latest estimates from UNAIDS (22) showing that countries of the first cluster (eastern/southern SSA) now have the lowest ECR of SSA, with an average value of 0.044 compared to values of 0.052 and 0.054 for countries of western/central SSA and countries of Sahel region respectively. An almost 20% lower value which, if sustained over time, is likely to lead to a more rapid control of the epidemic in those countries.

The effective contact rate has the advantage of being able to extract and convey information about the level of the HIV epidemics given the local context and lends itself more easily as a comparison tool. As public health programs have been implemented, the importance of the criteria against which these are evaluated has increased (12,43). While these metrics are critical in order to prioritize focus and optimize program and intervention implementations, they should only be used with caution to compare different HIV epidemics without first an extensive look at each local context. They also rely on national estimates of incidence and prevalence which may produce errors (44). The use of nationally aggregated data allows us to compare countries and clusters across SSA and are useful in the context of generalized epidemics, but are prone to ecological fallacy (45). They also overlook the salience of so-called high-risk key populations (46).

## Conclusion

While sociobehavioural factors play a key role, especially early on in nascent epidemics, latent indicators seem to play an important role in transmission dynamics and reduction of effective contact rates. Our use of principal component analysis and clustering techniques allowed us to identify the complex ways in which sociobehavioural characteristics evolved across SSA as the HIV epidemics progressed.

Our methods and findings can help guide further research, especially for targeted country-specific interventions in the pursuit of epidemic control. It should be noted that with the trend of HIV epidemics shifting from generalized epidemics to epidemics of smaller key populations, a people-centered approach is of the utmost importance to avoid risking stigmatization.

## Data Availability

This study used only publicly accessible aggregate-level data. All data produced in the present study are available upon reasonable request to the authors.

## References

1. Joint United Nations Programme on HIV/AIDS (UNAIDS). UNAIDS DATA 2021 [Internet]. 2021 [cited 2023 Feb 16]. Available from: https://www.unaids.org/sites/default/files/media_asset/JC3032_AIDS_Data_book_2021_En.pdf

2. Lee BX, Kjaerulf F, Turner S, Cohen L, Donnelly PD, Muggah R, et al. Transforming Our World: Implementing the 2030 Agenda Through Sustainable Development Goal Indicators. J Public Health Policy. 2016 Sep 1;37(1):13–31.

3. Brockerhoff M, Biddlecom AE. Migration, Sexual Behavior and the Risk of HIV in Kenya. Int Migr Rev. 1999;33(4):833–56.

4. Crampin AC, Glynn JR, Ngwira BM, Mwaungulu FD, Pönnighaus JM, Warndorff DK, et al. Trends and measurement of HIV prevalence in northern Malawi. AIDS [Internet]. 2003;17(12). Available from: https://journals.lww.com/aidsonline/Fulltext/2003/08150/Trends_and_measurement_of_HIV_prevalence_in.11.aspx

5. Guimarães MDC, Kendall C, Magno L, Rocha GM, Knauth DR, Leal AF, et al. Comparing HIV risk-related behaviors between 2 RDS national samples of MSM in Brazil, 2009 and 2016. Medicine (Baltimore). 2018 May 25;97(1 Suppl):S62–8.

6. Wirtz AL, Jumbe V, Trapence G, Kamba D, Umar E, Ketende S, et al. HIV among men who have sex with men in Malawi: elucidating HIV prevalence and correlates of infection to inform HIV prevention. J Int AIDS Soc. 2013;16(4S3):18742.

7. Hajizadeh M, Sia D, Heymann S, Nandi A. Socioeconomic inequalities in HIV/AIDS prevalence in sub-Saharan African countries: evidence from the Demographic Health Surveys. Int J Equity Health. 2014;13(1):18.

8. Annor FB, Chiang LF, Oluoch PR, Mang’oli V, Mogaka M, Mwangi M, et al. Changes in prevalence of violence and risk factors for violence and HIV among children and young people in Kenya: a comparison of the 2010 and 2019 Kenya Violence Against Children and Youth Surveys. Lancet Glob Health. 2022 Jan;10(1):e124–33.

9. Howard AL, Pals S, Walker B, Benevides R, Massetti GM, Oluoch RP, et al. Forced Sexual Initiation and Early Sexual Debut and Associated Risk Factors and Health Problems Among Adolescent Girls and Young Women -Violence Against Children and Youth Surveys, Nine PEPFAR Countries, 2007-2018. MMWR Morb Mortal Wkly Rep. 2021 Nov 26;70(47):1629–34.

10. Kaaya S, Siril H, McAdam K, Ainebyona D, Somba M, McAdam E, et al. Agents of change: Comparing HIV-related risk behavior of people attending ART clinics in Dar es Salaam with members of their social networks. PloS One. 2020;15(9):e0238240.

11. Merzouki A, Estill J, Orel E, Tal K, Keiser O. Clusters of sub-Saharan African countries based on sociobehavioural characteristics and associated HIV incidence. PeerJ. 2021 Jan;9:e10660.

12. Galvani AP, Pandey A, Fitzpatrick MC, Medlock J, Gray GE. Defining control of HIV epidemics. Lancet HIV. 2018;5(11):e667–70.

13. Jones A, Cremin I, Abdullah F, Idoko J, Cherutich P, Kilonzo N, et al. Transformation of HIV from pandemic to low-endemic levels: a public health approach to combination prevention. The Lancet. 2014;384(9939):272–9.

14. USAID. Quality information to plan, monitor, and improve population, health, and nutrition progrems.Estimates methods. 2019; Available from: https://dhsprogram.com/

15. Curtis SL, Sutherland EG. Measuring sexual behaviour in the era of HIV/AIDS: the experience of Demographic and Health Surveys and similar enquiries. Sex Transm Infect. 2004;80(suppl 2):ii22–7.

16. USAID. STATcompiler [Internet]. 2020. Available from: https://www.statcompiler.com/en

17. IHME. Sub-Saharan Africa Male Circumcision Geospatial Estimates 2000-2017 [Internet]. Institute for Health Metrics and Evaluation (IHME); 2020. Available from: http://ghdx.healthdata.org/record/ihme-data/sub-saharan-africa-male-circumcision-geospatial-estimates-2000-2017

18. World Bank. Rural population (% of total population) [Internet]. Washington, DC: The World Bank; 2020 May [cited 2020 Jun 15]. Available from: https://data.worldbank.org/indicator/SP.RUR.TOTL.ZS

19. World Bank. Gini index (World Bank estimate) [Internet]. Washington, DC: The World Bank; 2020 May [cited 2020 Jun 15]. Available from: https://data.worldbank.org/indicator/SI.POV.GINI

20. World Bank. tAntiretroviral therapy coverage (% of people living with HIV) [Internet]. Washington, DC: The World Bank; 2020 May [cited 2020 Jun 15]. Available from:https://data.worldbank.org/indicator/SH.HIV.ARTC.ZS

21. Zeev Maoz and Errol A. Henderson. The World Religion Dataset, 1945-2010: Logic, Estimates, and Trends. [Internet]. The World Bank; 2013 [cited 2020 Jun 15]. Available from: https://correlatesofwar.org/data-sets/world-religion-data/world-religion-data-v1-1

22. UNAIDS. 2020. AIDSinfo [Internet]. 2020. Available from: http://aidsinfo.unaids.org/

23. Pearson K. LIII. On lines and planes of closest fit to systems of points in space. Lond Edinb Dublin Philos Mag J Sci. 1901;2(11):559–72.

24. Lloyd S. Least squares quantization in PCM. IEEE Trans Inf Theory. 1982;28(2):129–37.

25. Petitjean F, Ketterlin A, Gançarski P. A Global Averaging Method for Dynamic Time Warping, with Applications to Clustering. Pattern Recogn. 2011 Mar;44(3):678–93.

26. Rousseeuw PJ. Silhouettes: A graphical aid to the interpretation and validation of cluster analysis. J Comput Appl Math. 1987;20:53–65.

27. Van Rossum G, Drake Jr FL. Python tutorial. Centrum voor Wiskunde en Informatica Amsterdam, The Netherlands; 1995.

28. McKinney W. Data Structures for Statistical Computing in Python. In: Walt S van der, Millman J, editors. Proceedings of the 9th Python in Science Conference. 2010. p. 51–6.

29. Harris CR, Millman KJ, van der Walt SJ, Gommers R, Virtanen P, Cournapeau D, et al. Array programming with NumPy. Nature. 2020;585:357–62.

30. Virtanen P, Gommers R, Oliphant TE, Haberland M, Reddy T, Cournapeau D, et al. SciPy 1.0: Fundamental Algorithms for Scientific Computing in Python. Nat Methods. 2020;17:261–72.

31. Tavenard R, Faouzi J, Vandewiele G, Divo F, Androz G, Holtz C, et al. Tslearn, A Machine Learning Toolkit for Time Series Data. J Mach Learn Res. 2020;21(118):1–6.

32. Pedregosa F, Varoquaux G, Gramfort A, Michel V, Thirion B, Grisel O, et al. Scikit-learn: Machine Learning in Python. J Mach Learn Res. 2011;12:2825–30.

33. Inc PT. Collaborative data science. 2015; Available from: https://plot.ly

34. Koopman JS, Jacquez JA, Welch GW, Simon CP, Foxman B, Pollock SM, et al. The Role of Early HIV Infection in the Spread of HIV Through Populations. JAIDS J Acquir Immune Defic Syndr [Internet]. 1997;14(3). Available from: https://journals.lww.com/jaids/Fulltext/1997/03010/The_Role_of_Early_HIV_Infection_in_the_Spread_of.9.aspx

35. Bailey RC, Moses S, Parker CB, Agot K, Maclean I, Krieger JN, et al. Male circumcision for HIV prevention in young men in Kisumu, Kenya: a randomised controlled trial. 2007;369:14.

36. Lei J hao, Liu L ren, Wei Q, Yan S bing, Yang L, Song T run, et al. Circumcision Status and Risk of HIV Acquisition during Heterosexual Intercourse for Both Males and Females: A Meta-Analysis. Tang JW, editor. PLOS ONE. 2015 May 5;10(5):e0125436.

37. Sharma SC, Raison N, Khan S, Shabbir M, Dasgupta P, Ahmed K. Male circumcision for the prevention of human immunodeficiency virus (HIV) acquisition: a meta-analysis. BJU Int. 2018;121(4):515–26.

38. Reed JB, Njeuhmeli E, Thomas AG, Bacon MC, Bailey R, Cherutich P, et al. Voluntary Medical Male Circumcision: An HIV Prevention Priority for PEPFAR. JAIDS J Acquir Immune Defic Syndr [Internet]. 2012;60. Available from: https://journals.lww.com/jaids/Fulltext/2012/08153/Voluntary_Medical_Male_CircumcisionAn_HIV.7.aspx

39. Mills EJ, Singh S, Nelson BD, Nachega JB. The impact of conflict on HIV/AIDS in sub-Saharan Africa. Int J STD AIDS. 2006 Nov;17(11):713–7.

40. Premkumar R, Tebandeke A. Political and socio-economic instability: does it have a role in the HIV/AIDS epidemic in sub-Saharan Africa? SAHARA-J J Soc Asp HIVAIDS. 2011 Jun 1;8(2):65–73.

41. Okware SI. Towards a national AIDS-control program in Uganda. West J Med. 1987 Dec;147(6):726–9.

42. Tanser F, Bärnighausen T, Grapsa E, Zaidi J, Newell ML. High Coverage of ART Associated with Decline in Risk of HIV Acquisition in Rural KwaZulu-Natal, South Africa. Science. 2013;339(6122):966– 71.

43. Ghys PD, Williams BG, Over M, Hallett TB, Godfrey-Faussett P. Epidemiological metrics and benchmarks for a transition in the HIV epidemic. PLOS Med. 2018 Oct;15(10):1–10.

44. Nsanzimana S, Remera E, Kanters S, Mulindabigwi A, Suthar AB, Uwizihiwe JP, et al. Household survey of HIV incidence in Rwanda: a national observational cohort study. Lancet HIV. 2017 Oct;4(10):e457–64.

45. Levin KA. Study Design VI -Ecological Studies. Evid Based Dent. 2006 Dec 24;7:108.

46. Barr D, P Garnett G, Mayer KH, Morrison M. Key populations are the future of the African HIV/AIDS pandemic. J Int AIDS Soc. 2021;24(S3):e25750.

