## Supplementary Material for "Changes of sociobehavioural characteristics and HIV in 29 Sub-Saharan African countries, 2000-2018"

#### Variables

| Attribute | Topic | Variable | Stratification | Categories |
|---|---|---|---|---|
| 1 | Demographic | Age under 25 | Men | |
| 2 | | | Women | |
| 3 | | Rurality | Men | |
| 4 | | | Women | |
| 5 | | Religion | | Christian |
| 6 | | | | Muslim |
| 7 | | | | Folk/Popular |
| 8 | | | | Unaffiliated |
| 9 | | | | Others |
| 10 | | Married or in union | Men | |
| 11 | | | Women | |
| 12 | | Number of wives (for men) and co-wives (for women) | Men | 1[1] |
| 13 | | | | ≥2[2] |
| 14 | | | Women | 0[3] |
| 15 | | | | 1[4] |
| 16 | | | | ≥2[5] |
| 17 | | Female headed household | | |
| 18 | | Literacy | Men | |
| 19 | | | Women | |
| 20 | | Access to media at least once a week | Men | |
| 21 | | | Women | |

[1] Percentage of currently married or in union men who have one wife

[2] Percentage of currently married or in union men who have two or more wives
[3] Percentage of currently married or in union women whose husband has no other wives
[4] Percentage of currently married or in union women whose husband has one other wife
[5] Percentage of currently married or in union women whose husband has two or more wives

| | | | |
|---|---|---|---|
| 22 | Employment | Worked in the last 12 months and is currently working | Men |
| 23 | | | Women |
| 24 | Wealth | Gini coefficient[6] | |
| 25 | Sexual behaviour | First sex by age 15 | Men |
| 26 | | | Women |
| 27 | | General fertility rate[7] | Women |
| 28 | | Use of contraception | Women |
| 29 | | Woman is justified asking for condom if husband has a sexually transmitted infection (STI) | Men |
| 30 | | | Women |
| 31 | | Mean number of sexual partners in lifetime | Men |
| 32 | | | Women |
| 33 | | Unprotected higher risk sex | Men |
| 34 | | | Women |
| 35 | | Ever paid for sexual intercourse | Men |
| 36 | | Unprotected paid sexual intercourse | Men |
| 37 | Gender-based violence | Wife beating justified | Men |
| 38 | | | Women |
| 39 | Women empowerment | Married women participating in decision making[8] | |
| 40 | | Married women who disagree with all reason justifying wife beating[9] | |
| 41 | HIV/AIDS | Comprehensive correct knowledge about AIDS[10] | Men |
| 42 | | | Women |

| | | | |
|---|---|---|---|
| 43 | | Ever received an HIV test | Men |
| 44 | | | Women |
| 45 | | Male circumcision | |
| 46 | | ART* coverage 2015 | |
| 47 | Accepting attitudes toward PLWHA | Would buy vegetables from shopkeeper with AIDS | Men |
| 48 | | | Women |

*Table S1 - Socioeconomic and behavioural variables included in the analysis. *ART = antiretroviral therapy.*

[6] The Gini coefficient indicates the level of wealth concentration in a country.
[7] Average number of children currently being born to women of reproductive age in the three years preceding the survey, expressed per 100 women age 15-44.
[8] Percentage of currently married women age 15-49 who usually make all three specific decisions either alone or jointly with their husband for (1) own health care, (2) large household purchases, and (3) visits to family or relatives.
[9] Percentage of currently married women age 15-49 who disagree with all five specific reasons justifying wife-beating: (1) burning food, (2) arguing with husband, (3) going out without telling him, (4) refusing sexual intercourse with him and (5) neglecting children.
[10] Percentage of men and women who correctly identify the two major ways of preventing the sexual transmission of HIV (using condoms and limiting sex to one faithful, uninfected partner), who reject the two most common local misconceptions about HIV transmission (ex. AIDS cannot be transmitted by mosquito bites, and it cannot be transmitted by supernatural means), and who know that a healthy-looking person can have HIV.

| Variable code | Year group | min | median | max | std |
| --- | --- | --- | --- | --- | --- |
| ART | 2000-2004 | 0.0 | 0.0 | 3.0 | 0.9 |
| | 2015-2019 | 25.0 | 56.0 | 79.0 | 14.3 |
| Access.to.media.M | 2000-2004 | 2.4 | 12.2 | 29.8 | 8.8 |
| | 2015-2019 | 1.7 | 7.5 | 47.5 | 11.6 |
| Access.to.media.W | 2000-2004 | 0.5 | 4.6 | 20.6 | 5.4 |
| | 2015-2019 | 0.3 | 4.0 | 20.1 | 4.7 |
| Age | 2000-2004 | 38.5 | 43.7 | 48.5 | 2.5 |
| | 2015-2019 | 36.3 | 41.7 | 44.8 | 2.7 |
| Buy.from.shopkeeper.with.AIDS.M | 2000-2004 | 18.3 | 50.0 | 83.9 | 17.3 |
| | 2015-2019 | 31.6 | 65.4 | 92.1 | 20.7 |
| Buy.from.shopkeeper.with.AIDS.W | 2000-2004 | 19.6 | 41.2 | 66.6 | 14.8 |
| | 2015-2019 | 26.5 | 63.8 | 89.2 | 21.5 |
| Christian | 2000-2004 | 3.3 | 60.9 | 88.6 | 25.8 |
| | 2015-2019 | 2.4 | 80.6 | 97.6 | 30.0 |
| Ever.paid.for.sex | 2000-2004 | 2.2 | 5.0 | 24.6 | 6.9 |
| | 2015-2019 | 2.1 | 8.0 | 18.4 | 4.8 |
| Ever.receiving.HIV.test.M | 2000-2004 | 2.1 | 10.5 | 24.6 | 6.2 |
| | 2015-2019 | 14.4 | 49.1 | 80.8 | 24.1 |
| Ever.receiving.HIV.test.W | 2000-2004 | 2.1 | 8.4 | 30.6 | 7.9 |
| | 2015-2019 | 17.3 | 67.8 | 86.9 | 24.2 |
| Female.headed.household | 2000-2004 | 9.4 | 24.7 | 41.5 | 8.8 |
| | 2015-2019 | 17.4 | 30.3 | 40.6 | 6.1 |
| First.sex.by.15.M | 2000-2004 | 2.0 | 9.0 | 25.8 | 7.0 |
| | 2015-2019 | 1.5 | 5.0 | 22.5 | 6.3 |
| First.sex.by.15.W | 2000-2004 | 3.0 | 20.4 | 36.3 | 10.7 |
| | 2015-2019 | 2.4 | 17.1 | 28.4 | 7.5 |
| Folk.Religion | 2000-2004 | 4.2 | 10.7 | 37.4 | 9.5 |
| | 2015-2019 | 0.0 | 3.8 | 18.1 | 4.3 |

| | | | | | |
|---|---|---|---|---|---|
| General.fertility.rate | 2000-2004 | 12.1 | 19.2 | 24.2 | 3.4 |
| | 2015-2019 | 11.8 | 16.3 | 23.0 | 3.2 |
| Justified.condom.if.husband.has.STI.M | 2000-2004 | 54.3 | 79.8 | 91.5 | 8.3 |
| | 2015-2019 | 69.6 | 80.6 | 98.5 | 9.0 |
| Justified.condom.if.husband.has.STI.W | 2000-2004 | 35.7 | 70.9 | 90.7 | 15.9 |
| | 2015-2019 | 14.3 | 75.8 | 97.3 | 20.2 |
| Knowledge.about.AIDS.M | 2000-2004 | 13.3 | 32.1 | 51.6 | 9.6 |
| | 2015-2019 | 17.4 | 39.8 | 68.8 | 15.0 |
| Knowledge.about.AIDS.W | 2000-2004 | 7.4 | 22.5 | 36.1 | 8.8 |
| | 2015-2019 | 10.9 | 42.2 | 66.9 | 17.0 |
| Literate.M | 2000-2004 | 33.3 | 76.9 | 88.9 | 19.1 |
| | 2015-2019 | 46.6 | 81.8 | 94.2 | 14.0 |
| Literate.W | 2000-2004 | 12.1 | 57.8 | 94.9 | 26.0 |
| | 2015-2019 | 22.1 | 67.1 | 97.0 | 22.1 |
| Married.or.in.union.M | 2000-2004 | 32.1 | 50.8 | 62.2 | 7.9 |
| | 2015-2019 | 37.0 | 50.5 | 61.4 | 6.8 |
| Married.or.in.union.W | 2000-2004 | 38.6 | 67.2 | 83.5 | 11.4 |
| | 2015-2019 | 51.7 | 60.6 | 81.4 | 8.0 |
| Married.women.participating.in.decisions | 2000-2004 | 11.4 | 31.8 | 49.5 | 11.9 |
| | 2015-2019 | 13.2 | 62.8 | 75.8 | 20.7 |
| Married.women.who.disagree.with.wife.beating | 2000-2004 | 8.6 | 27.2 | 68.4 | 15.3 |
| | 2015-2019 | 18.0 | 47.9 | 80.9 | 17.1 |
| Mean.number.of.sexual.partners.M.Normalized | 2000-2004 | 0.08 | 0.3 | 0.6 | 0.2 |
| | 2015-2019 | 0.02 | 0.3 | 0.8 | 0.2 |
| Mean.number.of.sexual.partners.W.Normalized | 2000-2004 | 0.07 | 0.2 | 0.5 | 0.1 |
| | 2015-2019 | 0.03 | 0.2 | 0.8 | 0.2 |
| Men.circumcised | 2000-2004 | 10.5 | 90.3 | 97.5 | 35.1 |
| | 2015-2019 | 14.6 | 93.0 | 98.9 | 31.8 |
| Men.who.work | 2000-2004 | 32.1 | 67.0 | 86.5 | 14.3 |
| | 2015-2019 | 58.7 | 81.1 | 91.9 | 9.1 |

| | | | | | |
|---|---|---|---|---|---|
| Muslim | 2000-2004 | 0.05 | 18.74 | 89.73 | 24.7 |
| | 2015-2019 | 0.08 | 12.953 | 96.38 | 31.0 |
| Number.of.co.wives.0 | 2000-2004 | 51.6 | 69.5 | 87.7 | 11.7 |
| | 2015-2019 | 61.2 | 83.7 | 93.2 | 11.6 |
| Number.of.co.wives.1 | 2000-2004 | 8.1 | 14.7 | 31.7 | 8.4 |
| | 2015-2019 | 1.9 | 13.2 | 32.5 | 9.3 |
| Number.of.co.wives.2 | 2000-2004 | 2.4 | 6.8 | 18.0 | 4.8 |
| | 2015-2019 | 0.4 | 3.1 | 10.0 | 2.9 |
| Number.of.wives.1 | 2000-2004 | 73.3 | 89.0 | 96.0 | 7.2 |
| | 2015-2019 | 74.1 | 92.7 | 97.9 | 7.1 |
| Number.of.wives.2 | 2000-2004 | 3.9 | 11.0 | 26.7 | 7.2 |
| | 2015-2019 | 2.1 | 7.3 | 25.9 | 7.1 |
| Other.Religion | 2000-2004 | 0.02 | 1.2 | 7.7 | 2.0 |
| | 2015-2019 | 0.0 | 0.4 | 2.7 | 0.7 |
| Unaffiliated.Religion | 2000-2004 | 0.11 | 0.5 | 6.0 | 1.4 |
| | 2015-2019 | 0.0 | 0.455 | 7.9 | 2.7 |
| Unprotected.higher.risk.sex.M | 2000-2004 | 0.0 | 20.8 | 63.1 | 14.0 |
| | 2015-2019 | 5.2 | 13.0 | 27.7 | 7.6 |
| Unprotected.higher.risk.sex.W | 2000-2004 | 0.0 | 10.6 | 31.3 | 7.8 |
| | 2015-2019 | 2.6 | 7.5 | 22.9 | 6.5 |
| Unprotected.paid.sex | 2000-2004 | 0.1 | 0.8 | 9.3 | 2.3 |
| | 2015-2019 | 0.1 | 0.7 | 2.5 | 0.7 |
| Use.of.contraception | 2000-2004 | 5.9 | 20.1 | 37.8 | 9.7 |
| | 2015-2019 | 5.4 | 22.8 | 48.9 | 13.6 |
| Wealth.index.Gini | 2000-2004 | 30.0 | 42.8 | 63.3 | 6.9 |
| | 2015-2019 | 33.0 | 43.5 | 57.1 | 6.0 |
| Wife.beating.justified.M | 2000-2004 | 16.1 | 44.4 | 74.8 | 15.8 |
| | 2015-2019 | 12.5 | 27.6 | 49.4 | 11.4 |
| Wife.beating.justified.W | 2000-2004 | 28.2 | 63.3 | 88.8 | 16.4 |
| | 2015-2019 | 16.3 | 41.4 | 79.4 | 17.7 |

| | | | | | |
|---|---|---|---|---|---|
| Women.who.work | 2000-2004 | 32.7 | 58.4 | 86.3 | 15.8 |
| | 2015-2019 | 33.3 | 61.9 | 77.8 | 14.7 |
| rural | 2000-2004 | 21.1 | 70.9 | 85.3 | 16.5 |
| | 2015-2019 | 36.6 | 67.6 | 87.6 | 16.1 |

#### Missing data imputation

We used scikt-learn's (1) implementation of a multiple iterative chained equation (2) imputation technique (3) with an extra-trees regressor with 100 estimators, and bounded between 0 and 100 as we are working with percentages.

Earlier surveys had a larger amount of missing values than more recent surveys, meaning the missing variables were not missing completely at random (4). No country had all missing values for any particular indicator, i.e. every country had at least one non-missing value for each of the 46 indicators between 2000 and 2018, which substantially improved our imputation results.

Indicators that have three or more (over 80 surveys) missing values and that have related indicators in the DHS surveys are imputed individually. These indicators are:

- Wife beating justified [W/M]
- Knowledge about AIDS [W/M]
- Buy from shopkeeper with AIDS [W/M]
- Justified condom if husband has STI [W/M]
- Mean number of sexual partners [W/M]
- Married women participating in decisions
- Married women who disagree with wife beating
- Ever paid for sex

To impute Ever.paid.for.sex and Mean.number.of.sexual.partners, we used the following additional indicators related to risky sexual behaviours:

- Higher risk sex in the last year
- Condom use at last higher risk sex (with a non-marital, non-cohabiting partner)
- Higher-risk Sex (with multiple partners among all respondents)
- Condom use during higher-risk sex (with multiple partners)

To impute Knowledge.about.AIDS, we used the following additional indicators:

- Men/Women who have heard of HIV or AIDS
- Knowledge of HIV prevention methods
- Use of condoms (prompted)
- Only one partner (prompted)
- Composite of 2 components (prompted)
- No incorrect beliefs about AIDS
- Healthy-looking person can have the AIDS virus
- AIDS cannot be transmitted by mosquito bites
- AIDS cannot be transmitted by supernatural means

- Cannot become infected by sharing food with someone who has AIDS
- Composite of 3 components
- Comprehensive correct knowledge about AIDS
- Knowledge of MTCT
- Knowledge of MTCT risk reduction

To impute Buy.from.shopkeeper.with.AIDS, we used the following additional indicators related to stigma and accepting attitudes towards PLWHA:

- Willing to care for family member sick with AIDS
- Female HIV+ teacher but not sick should be allowed to teach
- Not secretive about family member's HIV status
- Composite of 4 components
- Adult support of education on condom use

To impute Wife.beating.justified, Justified.condom.if.husband.has.STI, Married.women.participating.in.decisions, and Married.women.who.disagree.with.wife.beating, we used the following additional indicators related to spousal relationship:

- Justified in refusing sex with her husband if he has sex with other women
- Justified in asking for condom if she knows that her husband has an STI
- Justified in refusing sex if she knows husband has sexually transmitted disease
- Wife beating justified for at least one specific reason
- Husband jealousy [7 indicators]
- Non-categorical missing variables

The remaining missing variables related to literacy, access to media, HIV testing, number of wives and co-wives, and unprotected paid sex, which had less than three missing values were imputed without additional indicators.

#### Principal Component Analysis (PCA)

We used PCA to transform the 46 original dimensions of each survey into a smaller subset of uncorrelated indices called principal components (PCs) along which the variation of data is maximized and information loss is minimized. As the PCs consist of a linear combination of the initial 46 dimensions, they can be interpreted in terms of the original demographic, socioeconomic, and behavioral characteristics. The first 2 PCs explain the most variance and are used to represent the axes on a 2-dimensional (2D) space to provide a visual perspective of similarity between the surveys.

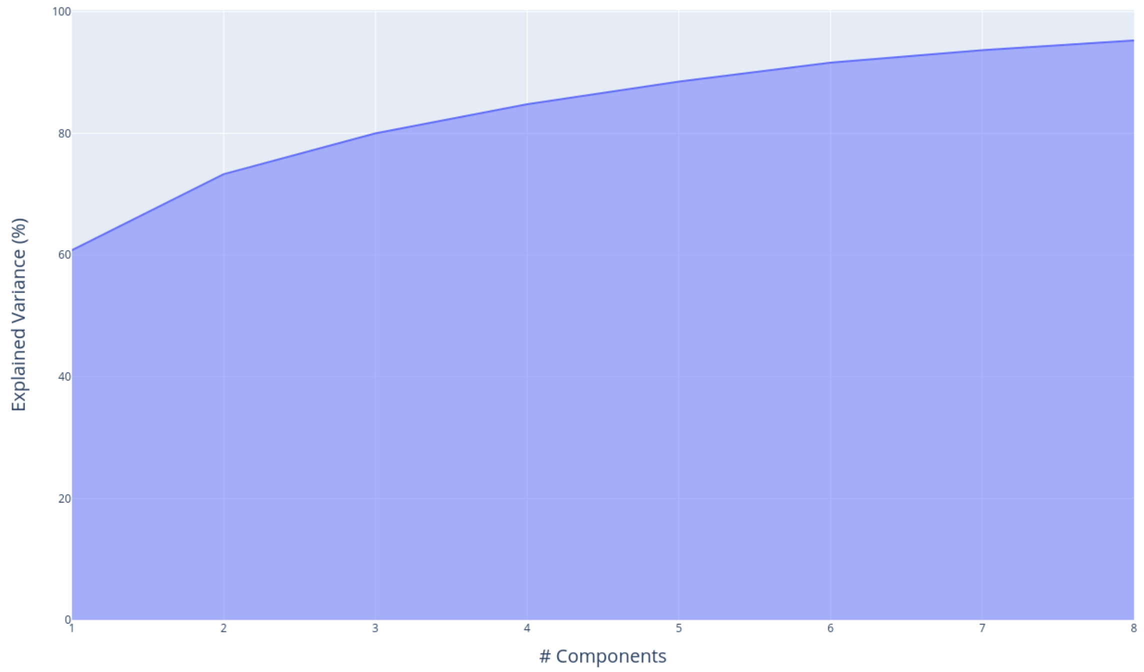

*Figure S1 Cumulative contribution of the PCs to total variance in the data set - the first 8 PCs account for over 95% of the variance.*

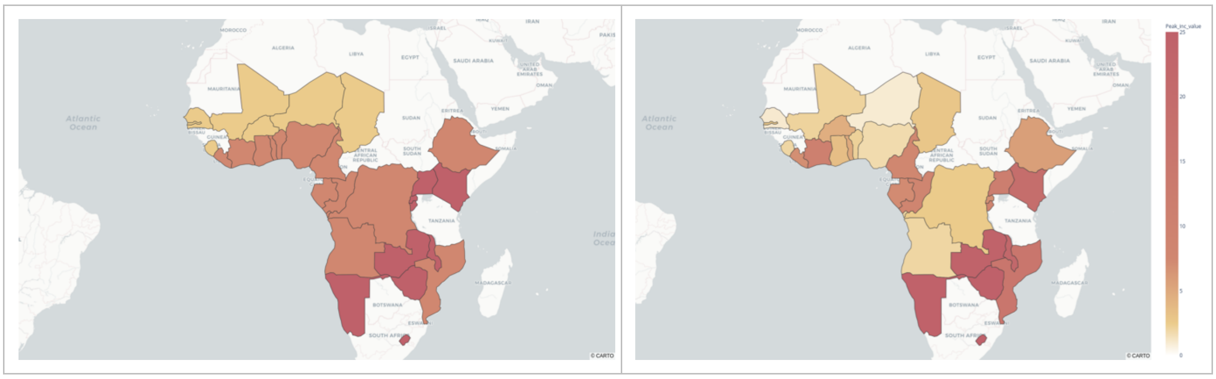

*Figure S2 - Visualization of sociobehavioural (SB) clusters across SSA. (left) Map of SSA showing SB clusters. (right) Map of SSA showing peak levels of HIV incidence.*

### Effective contact rate

In order to compare the progression of HIV epidemics across several SSA countries, we used the effective contact rate metric. An individual becomes exposed after an exposure event, from which it can become infected with the virus given a certain probability.

Let $r$ be the number of exposure events an individual has per year (this means number of drug injections, number of sexual partners, etc.), and let $\rho$ be the chance for a susceptible individual to contract the disease after such an exposure event.

$\rho$ is a generic factor which combines many into one, including:

- male circumcision

- condom usage
- other STDs which may increase chance of HIV infection
- PrEP usage
- ART coverage, if not already included above in r
- etc.

We can combine the two terms into one to give:

$$\beta = \rho * r$$

On a population-level however, the number of susceptible individuals ($S$) that will become infected and infectious ($I$) over the course of a year also depends on the proportion of $S$ itself in the population. Any factor impacting susceptibility of an individual (PrEP, condom use, etc.) is already factored into $\beta$.

The total number of new infections in a given year can thus be given by:

$$New\ infections[n] = \beta[n] * \frac{I[n]}{N[n]} * S[n]$$

where:

- S[n] is the number of susceptible in given year
- I[n] is the number of infectious in given year
- N[n] = S[n] + I[n] is the total population in given year
- β[n] is the effective contact rate

UNAIDS provides HIV incidence per 1000 population so:

$$Incidence[n] = New\ infections[n] * \frac{1000}{N[n]}$$

$$\Leftrightarrow Incidence[n] = \beta[n] * \frac{I[n]}{N[n]} * \frac{S[n]}{N[n]} * 1000$$

where:

- $\frac{I[n]}{N[n]} = Prevalence$ is the prevalence given by UNAIDS as a proportion (from 0 to 1)
- $\frac{S[n]}{N[n]} = 1 - \frac{I[n]}{N[n]} = 1 - Prevalence$ (from 0 to 1 depending on above)

We find that:

$$\beta = \frac{Incidence}{Prevalence * (1 - Prevalence)} * 10^{-3}$$

Estimates from UNAIDS of ART coverage, HIV incidence, and HIV prevalence are easily accessible and go back to 1990, and allow us to calculate the β, which can be interpreted as a proxy for all sociobehavioural characteristics.

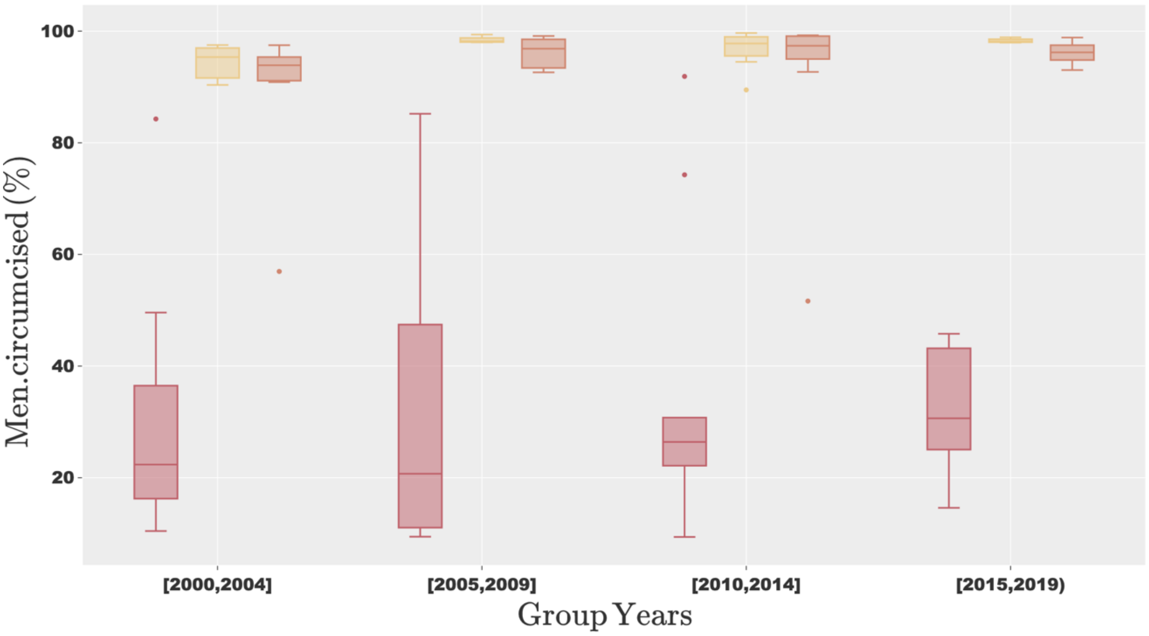

*Figure S3 – Progression of male circumcision per sociobehavioural cluster.*

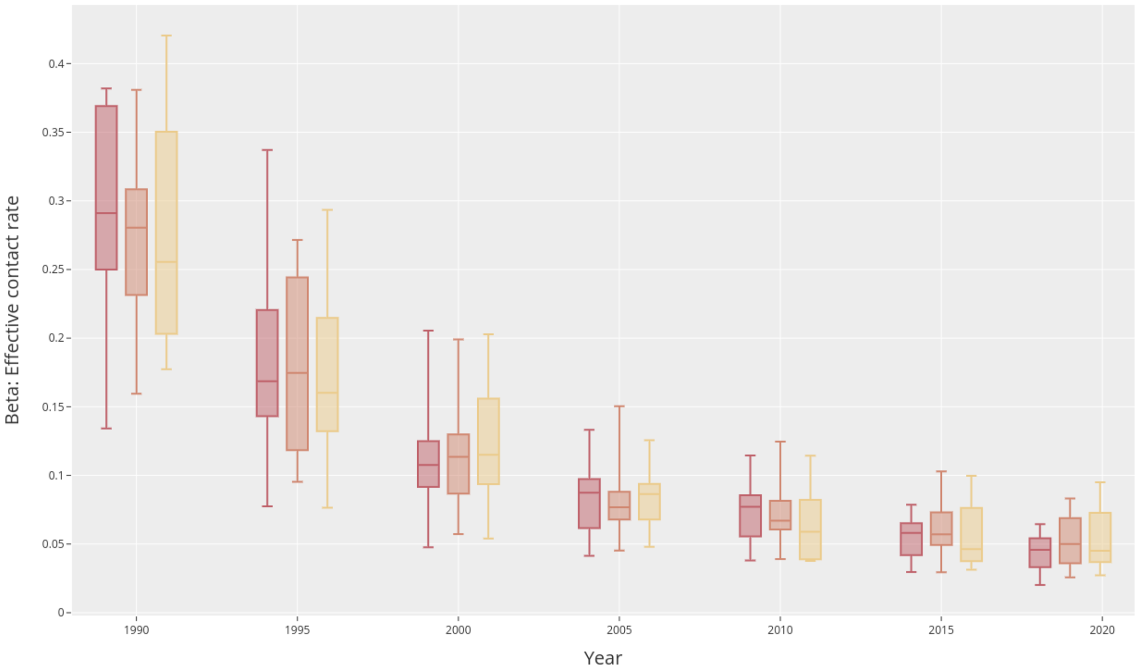

*Figure S4 - Progression of effective contact rates per sociobehavioural cluster.*

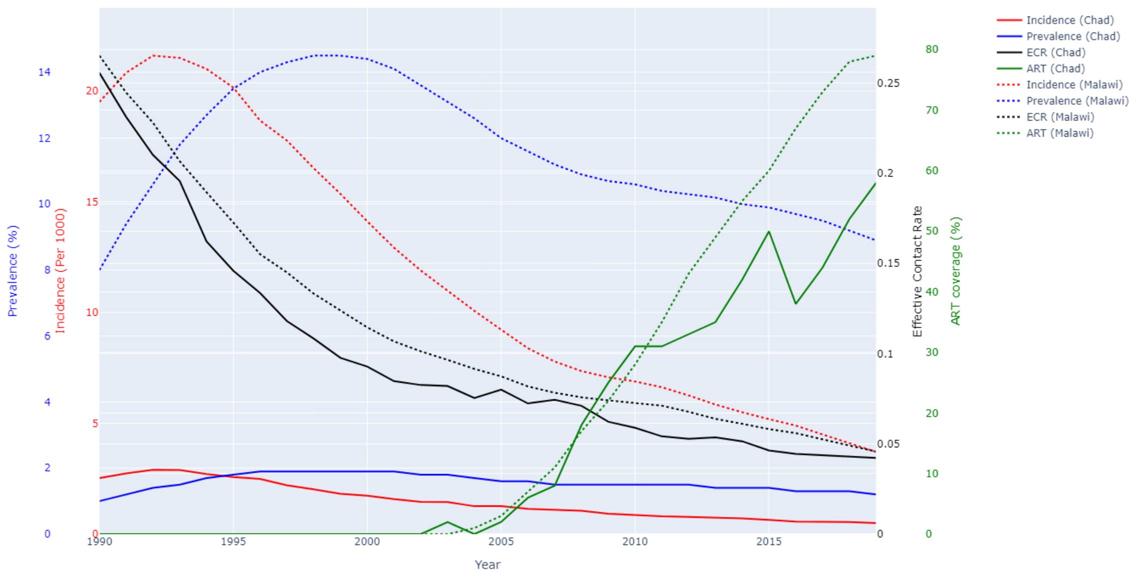

*Figure S5 – Progression of HIV prevalence, incidence, effective contact rate (ECR) and ART coverage from 1990 to 2018 in Chad and Malawi. Chad and Malawi have very different HIV epidemics (prevalence and incidence) but similar ECR evolutions.*

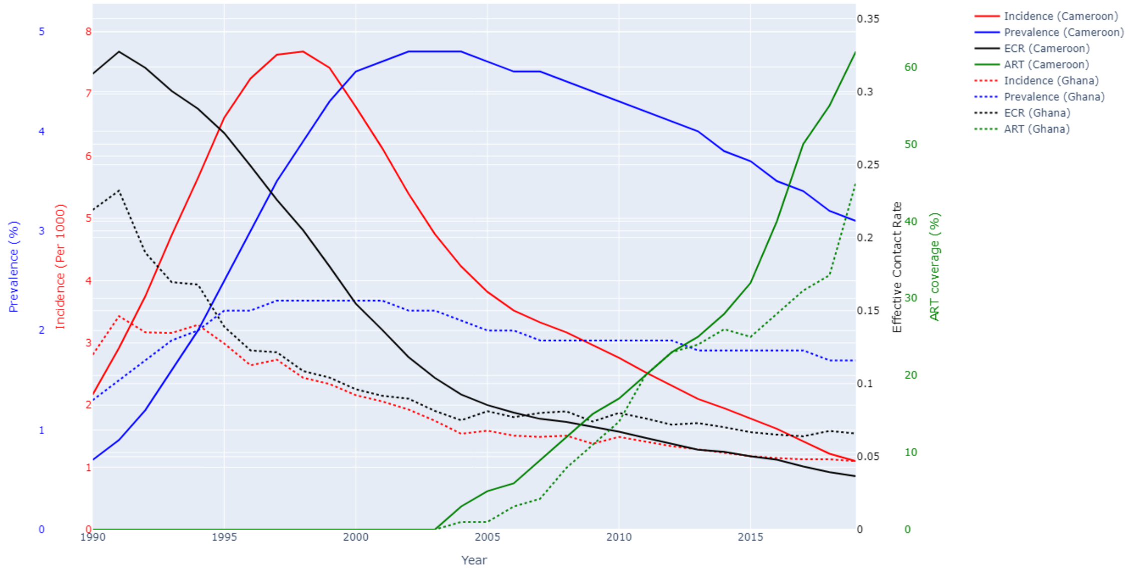

Figure S6 – Progression of HIV prevalence, incidence, effective contact rate (ECR) and ART coverage from 1990 to 2018 in Cameroon and Ghana. Cameroon and Ghana had similar epidemiological contexts in 1990 (incidence and prevalence) but have different evolutions of ECR. *The later inflection point on the curve of ECR in Cameroon is associated with a higher and later peak of HIV incidence.*

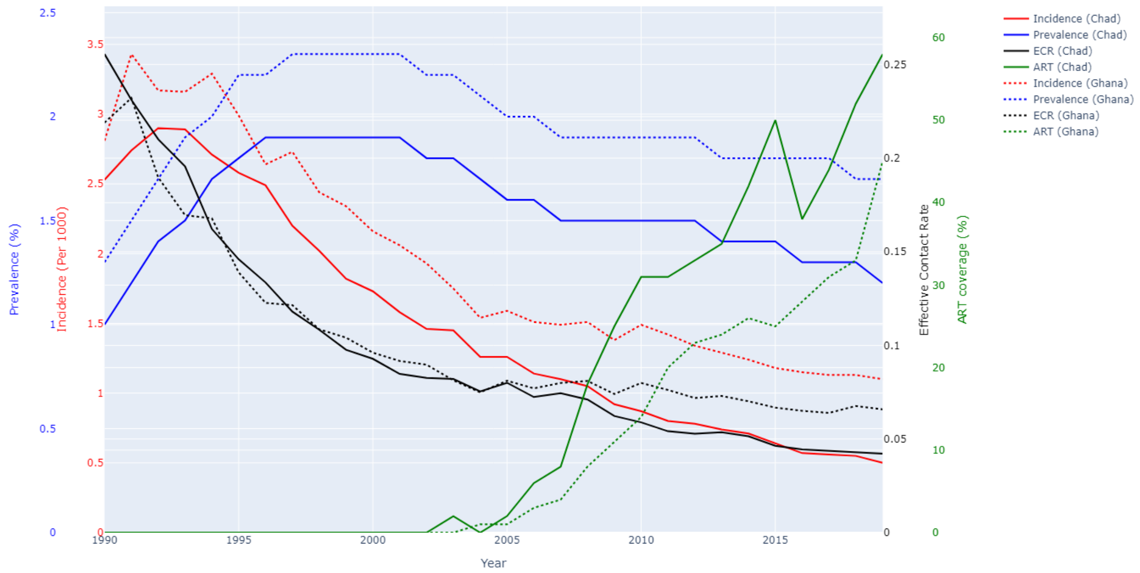

*Figure S7 - Progression of HIV prevalence, incidence and effective contact rate from 1990 to 2018 in Chad and Ghana. Chad and Ghana have both similar epidemiological contexts in 1990 (although they are in separate sociobehavioural clusters) and similar evolutions of ECR up to the year 2000. From 2000, Chad's effective contact rate decreases faster than Ghana, which associates with a faster increase in coverage of ART.*

### References

1. Pedregosa F, Varoquaux G, Gramfort A, Michel V, Thirion B, Grisel O, et al. Scikit-learn: Machine Learning in Python. J Mach Learn Res. 2011;12:2825–30.

2. Buuren S van, Groothuis-Oudshoorn K. **mice** : Multivariate Imputation by Chained Equations in *R*. J Stat Softw [Internet]. 2011 [cited 2019 Aug 19];45(3). Available from: http://www.jstatsoft.org/v45/i03/

3. Buck SF. A Method of Estimation of Missing Values in Multivariate Data Suitable for use with an Electronic Computer. J R Stat Soc Ser B Methodol. 1960;22(2):302–6.

4. Pedersen AB, Mikkelsen EM, Cronin-Fenton D, Kristensen NR, Pham TM, Pedersen L, et al. Missing data and multiple imputation in clinical epidemiological research. Clin Epidemiol. 2017 Mar 15;9:157–66.
